# Cortical activation predicts posttraumatic improvement in youth treated with TF-CBT or CCT

**DOI:** 10.1101/2022.06.03.22275968

**Authors:** Flint M. Espil, Stephanie Balters, Rihui Li, Bethany H. McCurdy, Hilit Kletter, Aaron Piccirilli, Judith A. Cohen, Carl F. Weems, Allan L. Reiss, Victor G. Carrion

## Abstract

**Background:** Identifying neural activation patterns that predict youths’ treatment response may aid in the development of imaging-based assessment of emotion dysregulation following trauma and foster tailored intervention. Changes in cortical hemodynamic activity measured with functional near-infrared spectroscopy (fNIRS) may provide a time and cost-effective option for such work. We examined youths’ PTSD symptom change following treatment and tested if previously identified activation patterns would predict treatment response.

**Methods:** Youth (*N* = 73, *mean age* = 12.97, *SD* = 3.09 years) were randomly assigned to trauma-focused cognitive behavioral therapy (TF-CBT), cue-centered therapy (CCT), or treatment as usual (TAU). Parents and youth reported on youth’s PTSD symptoms at pre-intervention, post-intervention, and follow-up. Neuroimaging data (*N* = 31) assessed at pre-intervention were obtained while youth engaged in an emotion expression task. Treatment response slopes were calculated for youth’s PTSD symptoms.

**Results:** Overall, PTSD symptoms decreased from pre-intervention through follow-up across conditions, with some evidence of relative benefit of TF-CBT and CCT over TAU but significant individual variation in treatment response. Cortical activation patterns were correlated with PTSD symptom improvement slopes (*r* = 0.53). In particular, cortical responses to fearful and neutral facial stimuli in six fNIRS channels in the bilateral dlPFC were important predictors of PTSD symptom improvement.

**Conclusions:** The use of fNIRS provides a method of monitoring and assessing cortical activation patterns in a relatively inexpensive and portable manner. Associations between functional activation and youths’ PTSD symptoms improvement may be a promising avenue for understanding emotion dysregulation in clinical populations.

## 1. Introduction

More than one fourth of youth within the United States will have been exposed to a traumatic event by late adolescence and more than a third will have experienced multiple traumatic events (Copeland et al., 2007; Copeland-Linder, 2008). Traumatic and adverse childhood experiences (TRACEs) are associated with many diverse outcomes and, if unaddressed, the impact may be enduring and last well into adulthood (Weems, Russell, Herringa, & Carrion., 2021). Childhood TRACEs increases the risk for a host of mental health conditions both in the short and long term including posttraumatic stress disorder (PTSD), anxiety, depression, attention deficit hyperactivity disorder (ADHD), suicidality, substance use, eating disorders, and psychosis (Alisic et al., 2014; Macdonald et al., 2010). Early developmental exposure to TRACEs also increases the risk for aggression, delinquent behavior, and violence in youth, these behaviors are closely linked to PTSD symptomatology (Aebi et al., 2017; Yoon et al., 2016). Childhood trauma may derail the normative trajectory of neurodevelopment resulting in impairments in executive functioning, memory, and learning (Carrion & Weems, 2017; Malarbi et al., 2017; Weems et al., 2021). Further, it can lead to structural and functional alterations in the brain and metabolic systems (Carrion & Weems, 2017; Shonkoff et al., 2012).

Many interventions have been developed to address the sequelae of childhood trauma ranging from individual, group, parent/family, school-based, and biological modalities. Core components of these interventions include trauma-informed child and parent assessment, stabilization and safety, psychoeducation about trauma responses, coping skills, emotional expression and regulation, cognitive processing, exposure to trauma reminders, consolidation of learned skills, and enhancement of self-efficacy and empowerment (Matlow, 2019). Evidence-based trauma-focused interventions are considered the first line of treatment for youth with posttraumatic stress (AACAP, 2010).

Of the evidence-based interventions for childhood trauma, trauma-focused cognitive behavioral therapy (TF-CBT) is the most widely researched (Cohen et al., 2017). Originally developed for use with sexual trauma, it has since been expanded to address a wide range of traumatic events including multiple traumatic experiences (Cohen et al., 2012). TF-CBT consists of nine PRACTICE components provided in three phases: 1) Stabalization Phase including psychoeducation, parenting, relaxation, affective modulation, and cognitive processing skills; 2) Trauma Narration and Processing phase, including trauma narration and processing; and 3) Integration Phase, including in-vivo mastery of traum reminders, conjoint child-parent sessions, and enhancing safety and future development. A wealth of studies demonstrates the efficacy of TF-CBT in reducing PTSD, depression, and other emotional and behavioral difficulties in traumatized youth (Cohen et al., 2017).

Cue-Centered Therapy (CCT) is a hybrid treatment combining elements of insight-oriented, CBT, psychodynamic, narrative, exposure, and family therapies (Carrion, 2016). It was designed to specifically address chronic trauma and the accumulation of stressors throughout the lifespan (i.e., allostatic load). The cue center focus, versus specific event focus, fosters intervention where multiple TRACEs are involved in precipitating emotional distress in youth. CCT includes caregiver participation in a few key sessions to minimize disruptions caused by barriers to caregiver attendance. CCT also aims to empower youth through facilitating insight into how traumatic cues develop through conditioning, and how these cues are linked to emotion, cognition, and behaviors, but also their physiology (Weissman et al., 2006). The four phases of CCT include: 1) psychoeducation and coping skills; 2) chronic traumatic stress history (i.e., trauma narratives and life timeline) and cognitive restructuring; 3) gradual exposure to trauma mimetic cues; 4) narrative retelling and consolidation of skills learned. A randomized controlled trial of youth with chronic trauma comparing CCT to a waitlist control group found that CCT was effective in reducing symptoms of PTSD, anxiety, and depression and improved overall functioning (Carrion et al., 2013). Caregivers in the CCT group also reported a significant reduction in anxiety. These gains were maintained at three-month follow-up.

Although interventions as TFCBT and CCT are effective there is variation in treatment response and evidence that different treatment modules may be relatively effective for different youth. Integrating the findings from neuroscience research among youth with TRACEs may help elucidate variability in treatment response (Weems & Neill, 2020). In particular, functional neuroimaging data suggest the potential of using brain biomarkers as objective assessment tools and even predictors of outcomes in youth with TRACEs (Carrion et al., 2008; Carrion et al., 2010; Garrett et al., 2012). Functional magnetic resonance imaging (fMRI) indicates that youth with TRACEs and PTSD may have differential activation and connectivity patterns in the amygdala, with concurrent involvement of the prefrontal cortex (i.e., medial, dorsolateral and ventrolateral prefrontal cortex; Herringa, 2017; Keding & Herringa, 2016; Wolf & Herringa, 2016). Frontocortical areas are vital for the appraisal of threatening stimuli and regulation of negative emotions (Etkin et al., 2011). Aberrant functioning (i.e., diminished PFC top-down control over amygdala) is thought to contribute to the heightened fear response in individuals with PTSD (Herringa et al., 2017; Koch et al., 2016; Sripada et al., 2012). The assessment of frontoamygdala activation and connectivity patterns allowed fMRI researchers to distinguish youth with TRACEs from those without (Garrett et al., 2012; Carrion et al., 2008; Keding & Herring,a 2016), and predict PTSD symptom improvement (Garrett et al., 2019; Cisler et al., 2016). Research suggests that establishing an understanding of the complex associations between functional activation in various brain areas and TRACEs among youth appears particularly promising for both research and clinical applications (Weems, Russell, Neill, & McCurdy, 2019; Weems et al., 2021; Weems & Neill, 2020). However, the high cost, motion sensitivity, and cumbersome environmental requirements of MRI hinder affordable and frequent assessment of these functional connectivity patterns.

In a first attempt to establish brain biomarkers of pediatric PTSD in an affordable and portable manner, Balters and colleagues (2021) examined functional activity among youth (n = 57) with TRACES and PTSD using a functional near-infrared spectroscopy (fNIRS) neuroimaging system. Hemodynamic responses were measured while participants engaged in a classic emotion expression task that included fearful and neutral face stimuli. A General Linear Model was applied to identify cortical activation associated with facial stimuli. Subsequently, a prediction model was established via a Support Vector Regression to determine whether PTSD severity could be predicted based on fNIRS-derived cortical response measures and individual demographic information. Results suggested increased activation in the dlPFC and vlPFC in response to negative emotion stimuli. Subsequent prediction analysis revealed ten features (i.e., cortical responses from eight frontocortical fNIRS channels, age, and sex) strongly correlated with PTSD severity. Examining if similar activation patterns and models predict treatment response would be an important next step in testing the utility of activation patterns to predict treatment response (Weems et al., 2019; Weems & Neill, 2020).

In this study, we conducted a randomized clinical trial comparing TF-CBT, CCT and Treatment as Usual (TAU) among youth in a community outpatient setting. We assessed participants at three time points: pre-intervention (T1; less than one week before intervention started), post-intervention (T2; less than one week after intervention concluded, and follow up (T3; three-months after conclusion of intervention). We hypothesized that 1)across conditions there would be treatment benefits that would vary by individual but that 2) CCT and TF-CBT would produce relatively better treatment outcomes than TAU, and 3) similar activation patterns found to predict PTSD symptom severity using fNIRS would also predict PTSD symptom treatment response.

## 2. Method

### 2.1 Participants and Procedure

This research was approved by the Stanford University Institutional Review Board and registered on clinicaltrials.gov (NCT02926677). Prior to participation parent consent and minor assent were obtained for all participants. All assessments, imaging, and therapy sessions were conducted within the Stanford Sierra Youth & Families community clinic in Sacramento, California, USA. Prospective study participants were recruited from treatment-seeking families referred through Sacramento County. County services comprise a network of five clinics serving families with MediCal insurance. All families referred to Stanford Sierra Youth & Families were initially reviewed during a standard intake process (medical and psychiatric history, semi structured interviews, parent- and child-report) by clinic staff for TRACEs and general study eligibility. Parents of youth ages 7-17 with TRACEs were subsequently connected with study staff to determine interest and conduct a formal screen for inclusion/exclusion criteria. Recruitment began in January 2016 and all participants concluded participation by July 2020.

All participants (*N* = 73) met the following inclusion criteria: (a) potentially traumatic event exposure and current PTSD symptoms as assessed by the UCLA PTSD Reaction Index-Parent and Child Report, (b) 7-17 years of age, (c) capacity to provide informed consent (parents) and assent (youth), (d) caregiver willing to participate in the study, (e) perpetrators of traumatic events not currently living with youth, (f) willingness to participate in fNIRS imaging, and (g) English fluency. Prospective participants were excluded for meeting any of the following: (e) currently taking antipsychotic or anti-seizure medication, (f) neurological disorders, (g) traumatic brain injury resulting in loss of consciousness, (h) major medical illness, (i) IQ < 70 as assessed by WASI-II (Wechsler, 2011), or (j) a comorbid diagnosis of psychotic disorder, bipolar disorder, severe substance abuse or dependence, acute suicidal ideation requiring hospitalization, or autism spectrum disorder.

Following enrollment, participants were evaluated at three time points: pre-intervention (T1), post-intervention (T2), and 3-month follow-up (T3). Both youth and parents completed self- and caregiver-measures, respectively, including structured interviews. Youth also completed fNIRS scanning and associated tasks at all three timepoints. To control for the effects of time, procedures were kept consistent across all participants in all three treatment groups. The CONSORT diagram (Figure 1) summarizes the flow of participants from referral through study completion and analysis. Of 247 youths referred, 102 were screened, 73 were enrolled and randomized, 67 attended at least one treatment session, 40 completed treatment and were assessed post-intervention, and 20 completed 3-month follow up assessment. Primary reasons for study drop out were losing contact with the research team, moving, deciding to pursue other care options, and being removed by the research team (e.g., due to substance use, cohabitating with the perpetrator of the traumatic event, primary medical issues, unable to attend study appointments due to traveling barriers, family requesting wrap-around services not provided in CCT or TF-CBT).

### 2.2 Materials

#### UCLA Child/Adolescent PTSD Reaction Index (UCLA PTSD-RI)

The UCLA PTSD-RI (Steinberg et al., 2013) is a self- and parent-report assessment of PTSD severity. Symptoms are rated across the three primary PTSD symptoms (i.e., intrusion, avoidance, and arousal) during the past month (0 = none to 4 = most of the time). Previous research on the PTSD-RI indicates excellent internal consistency across age ranges and racial/ethnic groups (alpha = .88-.91; Steinberg et al., 2013).

#### Multiaxial Anxiety Scale for Children, Second Edition (MASC)

The MASC (March et al., 1997) includes both youth self-report and parent-report versions to assess symptoms of anxiety disorders in youth. Scores are summed across four primary subscales (Physical, Harm, Avoidance, Social, Separation) to produce a total anxiety score. Previous research indicates acceptable levels of reliability and validity (Wei et al., 2014).

#### Child Depression Inventory (CDI)

The CDI (Kovacs, 2003) is a 27-item self-report questionnaire of depressive symptoms (0=absent, 1=mild, 2=severe) in youth ages 7-17. Items are summed for a total score ranging between 0 and 54 points, with higher scores indicating more severe depressive presentations. The CDI is used in both clinical and community settings, with previous studies indicating acceptable levels of reliability and validity (Sitarenios & Stein, 2004) of depressive symptoms in youth ages 8-19.

#### Wechsler Abbreviated Scale of Intelligence, Second Edition (WASI-2)

The WASI-2 (Wechsler, 2011) provides an estimation of overall intellectual ability (range 40-160) of individuals ages 6-89. In this study, we used the two-subtest (vocabulary and matrix reasoning) to estimate participants’ full scale IQ. Administration of the two-subtest form takes approximately 15 minutes, and research indicates acceptable levels of reliability and validity across studies (McCrimmon & Smith, 2012)

### 2.3 Interventions

#### *Trauma-Focused Cognitive Behavioral Therapy (*TF-CBT)

All TF-CBT therapists (Master’s level) completed initial TF-CBT training consisting of the 11-hour distance learning course TF-CBT *Web* (https://tfcbt2.musc.edu), a 2-day face-to-face training and bi-weekly consultation calls with a TF-CBT developer. During consultation calls, therapists received feedback on their current TF-CBT cases and discussed how to implement the model with appropriate fidelity for challenging situations. Fidelity was rated by a TF-CBT developer (author J.C.) who listened to audiotapes of all TF-CBT sessions. Specifically, fidelity was rated based on the following criteria: a) implementing the TF-CBT PRACTICE components in the appropriate order; over the appropriate length of time (i.e., 10-18 sessions); and with appropriate proportionality for each of the 3 phases of treatment; and b) completing at least 85% of required fidelity elements within each PRACTICE component for child and parent, respectively, using a detailed fidelity checklist. TF-CBT sessions occurred weekly and lasted 50 minutes in duration. All participants assigned to TF-CBT met for a minimum of 15 and maximum of 18 sessions depending on participant progress and availability.

#### *Cue-Centered Therapy (*CCT)

CCT therapists (Master’s level) completed a two day in-person CCT training. Therapists subsequently participated in weekly consult calls for the duration of the study with the CCT supervisor (author H.K.) to ensure treatment fidelity and address challenges. Additionally, all therapists completed one practice case before seeing study participants. All therapy sessions were recorded, and audio files were assessed for fidelity by the CCT supervisor. Fidelity was rated based on the following criteria: a) implementing CCT components in appropriate order; for the appropriate session and length of time (10-18 sessions) and b) completing at least 85% of the required fidelity elements for the treatment session, using a detailed fidelity checklist. Four additional CCT therapists were trained over the duration of the study to account for staff turnover. CCT sessions occurred weekly and lasted 50 minutes in duration. All participants assigned to CCT met for a minimum of 15 and maximum of 18 sessions depending on participant progress and availability.

#### Treatment As Usual (TAU)

Participants in the TAU group were seen for a similar number of sessions. Session content comprised supportive counseling, problem-solving, case management, and wrap-around services to meet the needs of youth and families. Therapists providing TAU were supervised by managers within the Stanford Sierra Youth & Families. Prior to participating in the study, all TAU therapists were vetted to ensure they had not been trained in either TF-CBT or CCT. Clinic managers also reviewed all progress notes written for services with study youth to assure services provided by TAU therapists did not include CCT or TF-CBT intervention elements. Consistent with the CCT and TF-CBT groups, youth receiving TAU met weekly, for 50 minutes, for 15-18 sessions.

### 2.4 Predictive fNIRS neuroimaging model

In a previous paper, we analyzed fNIRS neuroimaging data at T1 to predict PTSD symptom severity at T1 in this same sample (Balters et al., 2021). In the present study we reapplied our predictive model to assess whether it was possible to predict PTSD symptom improvement over the course of treatment (pre, post, and follow up) from fNIRS cortical signatures at T1. We present a methodological summary of the predictive fNIRS neuroimaging model below, and (see Balters et al., 2021 for additional details..

At T1, cortical hemodynamic activity of each participant was recorded over 36 measurement channels across bilateral dorsolateral, ventrolateral, orbitofrontal prefrontal regions (BA 8, 9, 10, 44, 45,46 and 47), and the right temporal and parietal regions (BA 21, 22 and 39) utilizing a continuous wave fNIRS system (NIRScout System, NIRx, Germany, ran at 7.81 Hz). Participants engaged in a facial expression task with fearful and neutral faces. A total of thirty fearful faces blocks, thirty neutral faces blocks, and thirty blank-screen blocks were presented in randomized order (2 second stimuli presentation, followed by 4-9 second jittered “rest period” black-screen). Participants determined the sex of the presented faces via a button press. Via NIRS Brain AnalyzIR Toolbox (Santosa et al., 2018) in Matlab 2016a (MathWorks Inc.), raw data were converted to optical density, band-pass filtered to eliminate systemic noise (4^th^ order Butterworth, 0.01-0.2 Hz), and corrected for motion artifacts using a wavelet-based method (Molavi and Dumont, 2012). The concentration changes of oxygenated hemoglobin (HbO) and deoxygenated hemo-globin (HbR) were computed according to the Modified Beer-Lambert (Wyatt et al., 1986), and noisy channels (∼10 %) were identified and removed using a correlation-based method (Cui et al., 2010).

### Data Analyses

#### Outcomes

Omnibus intervention outcomes were first tested with multilevel modeling analyses using HLM 7 (Raudenbush et al., 2011; see also Bryk & Raudenbush, 1987). The HLM analyses nested pre- to post- to follow-up change on Level 1 in outcome assessments as a function of treatment group, age, and gender on Level 2. These analyses began with examining overall trends in outcome assessment (Level 1 base models). HLM analyses use all available data – degrees of freedom reported below show variation in number of participants per outcome variable. We also made made planned contrasts between TAU versus TFCBT and TAU versus CCT with HLM and we analyzed the size of the effect of change within treatment groups separately for each time point. Specifically, we conducted paired samples *t*-tests and calculated Cohen’s *d* effect sizes to determine differences and magnitude of effect between pre and post, pre and follow up, and post and follow up within each treatment group - degrees of freedom show the number of participants for these analyses.

#### fNIRS predictive analyses

A General Linear Model (GLM) was applied to assess the task condition specific evoked hemodynamic responses. Based on the derived beta values, group-wise activation patterns for each task condition (fearful and neutral) were derived via a linear mixed effect model. One sample t-test (two-tailed) was then used to assess group-level cortical activations in response to different conditions (fearful/neutral face) and only HbO results were used for further analysis (*NOTE:* HbO measures are known to be more robust and sensitive to task-associated changes compared to HbR measures (Ferrari and Quaresima, 2012; Plichta et al., 2006)). Seventeen channels were tested to be significantly activated by fearful and neutral facial stimuli. The beta values of these channels were extracted from each participant as input for the predictive model, along with age and sex, resulting in a total of 19 input features. The predictive model consisted of a Pearson correlation coefficient-based feature selection strategy and cascaded support vector regression (Balters et al., 2021). The UCLA PTSD-RI score (i.e., averaged score of child and parent UCLA scores; Garrett et al., 2012; Garrett et al., 2019) were used as PTSD symptom severity score. Instead of training the model to predict UCLA PTSD-RI scores, in this study, the model was trained to predict PTSD symptom improvement. PTSD symptom improvement was captured via the trajectory of the UCLA PTSD-RI scores over time. To calculate change in symptom scores for the fNIRS analyses and in order to conduct similar analyses to our previous study, individual regression slopes were extracted across the time points (T1 to T2 to T3) using regression coefficient analysis (Lorch & Myers, 1990). Calculating regression coefficients differs from values derived from change scores in that regression coefficients are estimated per individual participant, where different observations are not independent from each other and these follow from but are distinct from the HLM approach used to test outcomes. Using individual regression slopes for each participant as their change in PTSD symptoms avoids the problems inherent in using difference scores or residual analyses. In this case, these producesd an “improvement” slope for each individual by creating a regression slope coefficient (Pfister et al., 2013). In this way the more negative a slope value the steeper the decrease in symptoms (e.g., a -3 would be better improvement than a -2 and a positive 3 would mean the symptoms were higher at the last assessment point). These valuses were then used as the criterion variable with the Pearson correlation coefficient-based feature selection strategy and cascaded support vector regression (Balters et al., 2021). In the current sample, 40 participants had at least two time points to calculate a slope and had either child or parent UCLA PTSD-RI reported symptoms or data from both parent and child. Additionally, we used average scores of the child and parent versions when both were available. Of these 40 participants, 31 also had valid fNIRS data.

## 3. Results

### 3.1 Demographic Information of the Participant Cohort

At T1, means and standard deviations were calculated for age and frequencies were calculated for sex, ethnicity, and race separately for each treatment group. TRACEs total represents the mean number of events experienced. Youth reported experiencing at least one of the following TRACEs: serious accidental injury, illness/medical trauma, community violence, domestic violence, school violence, physical assault, disaster, sexual abuse, sexual assault/rape, physical abuse, neglect, psychological maltreatment/emotional abuse, impaired caregiver, kidnapping/ abduction, bereavement, separation, forced displacement, trafficking/sexual exploitation, bullying, attempted suicide, witnessed suicide, and other trauma. Table 1 includes a description of the participant groups.

A total of 93% of participants reported experiencing multiple TRACEs while 4% reported experiencing one TRACE. The most common types of TRACEs were bereavement (74%), illness/medical trauma (71.2%), bullying (61.6%), domestic violence (58.9%), separation (56.2%), serious accidental injury (53.4%), school violence/emergency (38.4%), physical abuse (37%), and psychological maltreatment/emotional abuse (37%). Thirty-three percent of the sample have attempted suicide. Thirty-one children took psychotropic medication within the last 12 months and 16 children had previously taken psychiatric medication. Lifetime psychiatric comorbidities included ADHD, aggression/oppositional, panic, anxiety, bipolar, borderline personality disorder, bullying, complex PTSD, depression, incontinence, and suicidal ideation (with and without self-harm).

### 3.2 Preliminary and Dropout Analyses

There were no significant differences among treatment groups across age, gender, ethnicity, or average number of traumatic events (Table 1). There were no statistically significant baseline differences among groups on the child and parent PTSD-RI reports (Table 2). Chi-squared tests of independence and *t*-tests revealed no significant differences on baseline characteristics (age, race, ethnicity, clinical outcome measures) between those who dropped or remained in the study. Examination of score distributions indicated the planned analyses were appropriate.

### 3.3 Treatment Outcome Analyses

HLM analyses and examination of the means indicated differences in scores and significant decreasing trends using the Level 1 effect of time on UCLA PTSD-RI child report symptoms [slope = -6.47, S.E. = 1.28, *t*(55) = -5.03, *p* < .001], UCLA PTSD-RI parent report symptoms [slope = -3.61, S.E. = 1.28, *t*(56) = -2.82, *p* < .01], MASC anxiety symptoms [slope = -5.79, S.E. = 1.84, *t*(59) = -3.15, *p* < .01], and CDI depression [slope = -2.38, S.E. = 0.62, *t*(63) = -3.87, *p* < .001]. Overall, these results suggest improvement across each of the treatment outcomes. We did not find a differential effect of the three-treatment group on symptoms trajectories on level 2. For example, using the UCLA PTSD-RI child report symptoms, the results indicated a nonsignificant effect of treatment group on change [slope = -0.45, S.E. = 1.59, *t*(69) = -0.28, *p* = 0.78]. Similarly, using UCLA PTSD-RI parent report symptoms indicated a nonsignificant effect of treatment group on change [slope = -0.54, S.E. = 1.59, *t*(69) = -0.34, *p* = 0.73] as did the MASC anxiety symptoms [slope = -0.95, S.E. = 2.47, *t*(66) = -0.39, *p* = 0.70] and CDI depression [slope = 0.31, S.E. = 0.74, *t*(68) = 0.42, *p* = 0.67]. When entering age and gender into the specified model, we found a significant effect of age but not treatment group on UCLA PTSD-RI parent symptom reports [slope = 6.47, S.E. = 2.49, *t*(53) = 2.60, *p* < .05]. Graphing the effect (see Figure 2) revealed that males had a steeper downward slope over time compared to females. Additionally, it was found that there was a trending effect of age but not treatment group on CDI depression [slope = -0.39, S.E. = 0.23, *t*(60) = -1.66, *p* = 0.10], indicating a steeper downward slope for the upper quartile age group (average age = 17.12) compared to the lower quartile age group (average age = 8.84).

We next made planned contrasts between TAU versus TFCBT and TAU versus CCT. For TFCBT versus TAU contrast, there was not a significant effect of condition on change in UCLA PTSD-RI child report symptoms, MASC anxiety symptoms, or CDI depression; however, there was a trending effect of change on UCLA PTSD-RI parent report symptoms [slope = -5.61, S.E. = 2.82, *t*(35) = -1.99, *p* = 0.055] with relative improvement for the TFCBT group compared to TAU. The trending effect of TFCBT on PTSD-RI parent report symptoms is illustrated in Figure 2. For CCT versus TAU contrast, there was not a significant effect of condition on change in UCLA PTSD-RI child report symptoms, UCLA PTSD-RI parent report symptoms, MASC anxiety symptoms, or CDI depression.

Given the trends seen in the planned contrasts and the potential for relatively small sample sizes particularly at T3 may have obscured omniubus differences in the HLM analyses testing pre, post, and follow-up, we analyzed the size of the effect of change within treatment groups separately for each time point. Specifically, we conducted paired samples *t*-tests and calculated Cohen’s *d* effect sizes to determine differences and magnitude of effect between pre and post, pre and follow up, and post and follow up within each treatment group again degrees of freedom show the number of participants for these analyses. Because the paired *t*-tests indicated significant differences within the TFCBT and CCT treatment groups, we used Cohen’s *d* to compare within-subjects relative differences at each time point by treatment group (see Table 2)

#### UCLA PTSD-RI Child Report Symptoms

The TAU group showed no significant decreases in UCLA PTSD-RI child reported symptoms from pre to post, post to follow up, or pre to follow up. Within the TFCBT group, there was a significant decrease from pre to follow up [*t*(6) = 2.97, *p* < .05] with a large effect size (Cohen’s *d* = 1.12). For youth who received CCT, there was a significant decrease from pre to follow up [*t*(7) = 3.13, *p* < .05] with a large effect size (Cohen’s *d* = 1.11).

#### UCLA PTSD-RI Parent Report Symptoms

The TAU group showed no significant decreases in UCLA PTSD-RI parent reported symptoms from pre to post, post to follow up, or pre to follow up. Within the TFCBT group, there was a significant decrease from pre to follow up [*t*(6) = 2.86, *p* < .05] with a large effect size (Cohen’s *d* = 1.08). Within the CCT group, there was a significant decrease from post to follow up [*t*(7) = 3.20, *p* < .05] with a large effect size (Cohen’s *d* = 1.13). Additionally, there was a significant decrease from pre to follow up [*t*(7) = 2.85, *p* < .05] with a large effect size (Cohen’s *d* = 1.01).

#### MASC Symptoms

The TAU group showed no significant decreases in MASC symptoms from pre to post, post to follow up, or pre to follow up. Within the TFCBT group, there was a significant decrease from post to follow up [*t*(6) = 2.67, *p* < .05] with a large effect size (Cohen’s *d* = 1.01). Within TFCBT, there was also a significant decrease from pre to follow up [*t*(7) = 3.23, *p* < .05] with a large effect size (Cohen’s *d* = 1.14). For the CCT group, there was a significant decrease from pre to post [*t*(11) = 3.85, *p* < .01] with a large effect size (Cohen’s *d* = 1.11).

#### CDI Symptoms

There were no significant decreases in CDI scores for the TAU group, TFCBT group, or CCT group from pre to post, post to follow up, or pre to follow up.

### 3.4 Predictive fNIRS Neuroimaging Model for PTSD Symptom Improvement

The process of the predictive model training is illustrated in Figure 3, in which correlation coefficients, mean square error (MSE) and the ratio between correlation and MSE are presented across different feature sets (Figure 3A-C). The overall prediction performance of the model was assessed by the ratio of its Pearson’s correlation and mean square error (MSE).

Six features resulted in the optimal feature set as evidenced by the highest ratio value between correlation coefficient between the predicted scores and the target slope score (*r* = 0.53, *p* = .0028, Figure 3E) and MSE loss (black dash circle in Figure 3C). The six features were composed of cortical activation patterns within six channels (Figure 3D), including increased cortical activation in the left dlPFC (channels 9,10,11) in response to fearful facial stimuli, increase cortical activation in the left dlPFC (channel 2) in response to neutral facial stimuli, and increased cortical activation in the right dlPFC (channels 18,28) in response to neutral facial stimuli. Age and gender were not part of the optimal feature set.

To better understand the underlying mechanisms of the prediction model results, the associations between cortical activation and PTSD symptom improvement for each of the six channels were inspected (see Figure 4). Results showed statistically significant correlation between normalized beta values and PTSD symptom slope for channel 9 (*r* = 0.421, *p* = 0.018), channel 10 (*r* = 0.448, *p* = 0.006), and channel 28 (*r* = -0.403, *p* = 0.027).

## 4. Discussion

In this study we conducted a small randomized controlled trial to compare the effectiveness of three separate interventions on PTSD symptoms and examine the utility of fNIRS in predicting improvement among youth. We hypothesized that across conditions there would be treatment benefits that would vary by individual but that 1) TF-CBT and CCT would produce relatively better treatment outcomes than TAU. Consistent with hypothesis 1, youths who received CCT and TFCBT reported significantly lower levels of both PTSD and anxiety by follow-up while those in the TAU group indicated no significant differences over time. Additionally, there was no reduction in depressive symptoms in any of the three groups. With respect to hypothesis 2, the results showed that task-evoked response from six fNIRS channels within the dlPFC allow prediction of PTSD symptom improvement. These findings add to the emerging fNIRS literature demonstrating the feasibility of assessing cortical signatures of symptomatic improvement in an affordable and portable manner, and specifically underscores the utility of this approach for youth with posttraumatic symptoms.

The PTSD symptom reduction seen among youth receiving TF-CBT is consistent with previous studies. In a meta analysis of 21 randomized controlled trials of TF-CBT versus a waitlist control group or active treatment, Lenz and Hollenbaugh (2015) demonstrated large to moderate effect sizes, respectively. Additional analyses indicated those who receive TF-CBT also reported decreases in depressive symptoms, although these effect sizes were in the moderate to small ranges in waitlist control and active comparison treatments, respectively. Comparisons to previous research on CCT are less available, however. In the only other trial conducted to date, Carrion et al. (2013) found CCT was more effective than a waitlist control. The findings from the present study provide further support for the effectiveness of CCT, but future trials are necessary to continue developing the evidence for this approach.

Although both TF-CBT and CCT were effective in reducing PTSD symptoms, those in the TAU condition also reported some level of symptom reduction. This finding is consistent with previous research conducted in community-based settings such as outpatient clinics. For example, in a meta-analysis of 52 randomized controlled trials, Weisz (2013) and colleagues found lower effect sizes for evidence-based youth psychotherapies compared with usual care. This effect also became non-significant when accounting for how participants were enrolled (clinically referred vs youth presenting to clinics with diagnoses). In a large randomized effectiveness trial of evidence-based approaches for youth anxiety, depression, trauma, and conduct problems conducted by community clinicians from multiple outpatient clinics, the evidence-based psychotherapy approach was not superior to usual care on any clinical measure (Weisz et al., 2020). Consistent with these findings, the meta -analysis conducted by Lenz and Hollenbaugh (2015), effect sizes for PTSD symptom improvement were also smaller when compared against active, alternative treatments. These findings do not suggest that TF-CBT or CCT are no more effective than TAU, but that the lack of robust differences in our results is consistent with other studies conducted in community-based settings.

Results further demonstrated that PTSD symptom improvement could be predicted via fNIRS cortical signals. Specifically, results showed that the emotion-evoked response from six fNIRS channels in the bilateral dlPFC correlated with PTSD symptom improvement (*r* = 0.53).

These findings align with fNIRS and fMRI studies that associate the bilateral dlPFC with pediatric and adult PTSD (Balters et al., 2019; Fonzo et al., 2017; Garrett et al., 2012) and demonstrate associations between treatment-related PTSD symptom reduction and normalization of dlPFC brain function in adults (Matsuo et al., 2009; Fonzo et al., 2017). With respect to prediction performance, there is, to our knowledge, only one other study that has established association between fNIRS cortical signatures and PTSD symptom improvement (Matsuo et al., 2009). The study focused on adults with PTSD and found a significant positive association between the longitudinal change in cortical activation (i.e., difference between pre- and post-treatment) in the lateral PFC and PTSD symptom improvement (*r* = 0.66, *p* = 0.02, *N* = 12). A likely reason for the reduced prediction performance in this study could be that the present data was collected at mid-treatment and therewith potentially decreased the effect of symptom improvement. An additional explanation could be that Matsuo et al. (2009) used recall events as stimuli which could potentially result in increased fear responses compared to our facial expression task (Miller et al., 2021). Lastly, Matsuo et al. (2009) used an adult sample, and our youth sample could contain more variance in cortical responses that are associated with brain development (Keding & Herringa, 2016; Herringa, 2017). Future research should consider these experimental variables and create additional benchmarks of PTSD cortical signatures in youth. Overall, our findings suggest that there are associations between fNIRS cortical signatures and PTSD symptom improvement in youth, and that task-related fNIRS assessments are a promising direction for the development of effective prediction models of PTSD symptom improvement.

While the predictive model derived the optimal feature set to best predict PTSD symptom severity on the given dataset, the model does not provide information about the underlying cortical activation pattern. The optimal feature set could therefore represent a combination of cortical activation pattern that could include both increased and decreased responses to the facial stimuli. To better understand the underlying cortical mechanisms of the prediction model results, the associations between cortical activation and PTSD symptom improvement for each of the six channels were inspected. Results showed overall positive correlations between the activation of the channels in left dlPFC (i.e., channels 2,9,10,11) during fearful and neutral faces and PTSD symptom slopes. In other words, higher symptom improvement (i.e., more negative PTSD symptom slopes) was associated with lower cortical responses to fearful and neutral faces. These findings align with previous studies that identified dysregulated brain functioning during negative emotion processing and regulation in the left dlPFC in association with adult PTSD (Fonzo et al., 2017). The dlPFC is part of a prefrontal circuitry that is known to be involved in emotion regulation (i.e., top-down control over amygdala; Ganella et al., 2017; Gee, 2016). Reduced activation of the left dlPFC in individuals with higher PTSD symptom improvement therefore suggest that these individuals had a more normalized threat response (i.e., diminished activation of the frontoamygdala circuity to facial stimuli). In this context it is important to note that there is increased evidence that youth with PTSD, in contrast to adults with PTSD, exhibit fear responses not only to fearful faces but also neutral faces (Garrett et al., 2012; Garrett et al., 2019; Keding and Herringa, 2016; Balters et al., 2021). Underlying reason for the fear response has been attributed to the ambiguity of neutral faces that could be potentially perceived as threatening to youth with PTSD (Garrett et al., 2019). Our findings regarding the left dlPFC, thus, indicate an association between normalization of brain function within this region and PTSD symptom improvement.

In contrast to the positive association between PTSD symptom slope and activation in the left dlPFC, findings are overall indicative of negative associations between the right dlPFC and PTSD symptom slopes in response to neutral faces. In other words, increased symptom improvement (i.e., more negative PTSD symptom slope) was associated with higher activation in the right dlPFC. The right dlPFC is known to be involved in cognitive attention control (Kondo et al., 2004; Sanchez-Lopez, 2018) and researchers have argued for the causal role of the right dlPFC in the generation of attentional impairments that are implicated in emotional disturbances such anxiety (Sanchez et al., 2016). Because “problems in concentration” is a common DMS-5 symptom (American Psychiatric Association, 2013) in youth with PTSD symptoms (Russel et al., 2017), our findings suggest that PTSD symptom improvement leads to normalization of cortical function in the right dlPFC. Individuals with higher levels of PTSD symptom improvement utilized more cognitive resources in the right dlPFC that potentially afforded them with higher attentional control (i.e., higher levels of concentration). In the context of pediatric PTSD symptomatology our results show one more interesting finding. In our previous study that utilized fNIRS to predict PTSD symptom severity, the right middle temporal gyrus (MTG) was identified as an important prediction feature (Balters et al., 2021). In our present study, the MTG was not detected as an important prediction feature. Activation in the right MTG has been associated with dissociation, another key symptom in PTSD (Lanius et al., 2002; 2005). The absence of the MTG as important prediction feature of PTSD symptom improvement is a further sign of normalization of aberrant cortical brain function in pediatric PTSD. Lastly, in contrast to the previous findings that also identified age and sex as important prediction features of PTSD symptom severity, the present findings did not determine age and sex as an important prediction feature. While the underlying cause for the absence of age and sex as prediction features could be the reduced sample size in this study (n = 31 vs. n = 56), the results could also suggest that PTSD-related alternations in brain functioning are sex- and age dependent, and normalize with symptom improvement. Future research needs to be conducted to systematically study the effect of age and gender in the context of pediatric PTSD symptom improvement.

Although these data provide a foray into the utility of fNIRS to measure and predict treatment outcomes among youth with history of TRACEs and PTSD symptoms, there are several important limitations. The small sample size limited statistical power in our larger predictive model and treatment outcome comparisons among groups. Premature termination in youth mental health care ranges from 28%-75% and is even higher in effectiveness than in efficacy studies (de Haan, Boon, de Jong, Hoeve, & Vermeiren, 2013). Although common in effectiveness trials conducting treatment research in community mental health settings (e.g., Chorpita et al., 2015; Weisz et al., 2020), loss of data due to drop out at post-intervention and follow-up lowered our statistical power and ability to draw conclusions about treatment outcomes. Future studies examining cortical activation using fNIRS among youth with TRACEs should incorporate larger samples to replicate these findings. Additionally, concomitant fNIRS and fMRI imaging could be conducted to corroborate fNIRS findings and further elucidate how cortical activity measured during fNIRS corresponds to activity in subcortical areas. Future work should also incorporate age and sex-matched healthy controls (i.e., youth without a history of TRACEs) to examine how patterns of activation measured using fNIRS may differ before and after treatment.

Despite these limitations, our study was the first to use fNIRS to examine changes in cortical activation among youth with history of TRACEs receiving evidence-based approaches to treat PTSD symptoms. Given the barriers to traditional (i.e., fMRI) imaging methods, fNIRS provides a cost-effective, flexible method for measuring cortical activity. We also conducted fNIRS imaging within a community clinic among typical treatment seeking families. This approach creates opportunities to collect functional imaging data among a more ecologically valid treatment seeking sample compared to traditional (e.g., university hospital, academic medical center) settings. Our sample was highly heterogeneous and diverse, which allowed for greater external validity in interpreting results. Lastly, the differential improvement within TF-CBT and CCT also provides further support for these interventions for youth with PTSD symptoms.

## Data Availability

All unidentifiable data produced in the present study are available upon reasonable request to the authors

## Appendix

**Table 1.** Demographics for Control versus Treatment Groups

|  | TAU | TF-CBT | CCT |
| --- | --- | --- | --- |
| Number of participants | 26 | 22 | 25 |
| Age | M=13.04<br>(SD=3.05) | M=13.41 (SD=3.30) | M=12.52 (SD=3.00) |
| Sex |  |  |  |
| *Female* | 61.5% | 59.1% | 68% |
| *Male* | 38.5% | 40.9% | 32% |
| Ethnicity |  |  |  |
| *Hispanic/Latino* | 11.5% | 31.8% | 28% |
| *Not Hispanic/Latino* | 53.8% | 50% | 52% |
| *Missing* | 34.6% | 18.2% | 20% |
| Race |  |  |  |
| *African American* | 26.9% | 27.3% | 20% |
| *Asian* | 7.7% | 0% | 0% |
| *American Indian/Alaskan* | 3.8% | 0% | 12% |
| *Pacific Islander* | 0% | 4.5% | 0% |
| *White* | 57.7% | 50% | 60% |
| *Other* | 7.7% | 18.2% | 12% |
| TRACEs Total (T1) | M=7.73 (SD=3.44) | M=7.10 (SD=3.19) | M=6.96 (SD=3.52) |

*Note.* We use the following abbreviations: mean (M), standard variation (SD), Treatment As Usual (TAU), Trauma-Focused Cognitive Behavioral Therapy (TF-CBT), Cue-Centered Therapy (CCT).

**Table 2.**
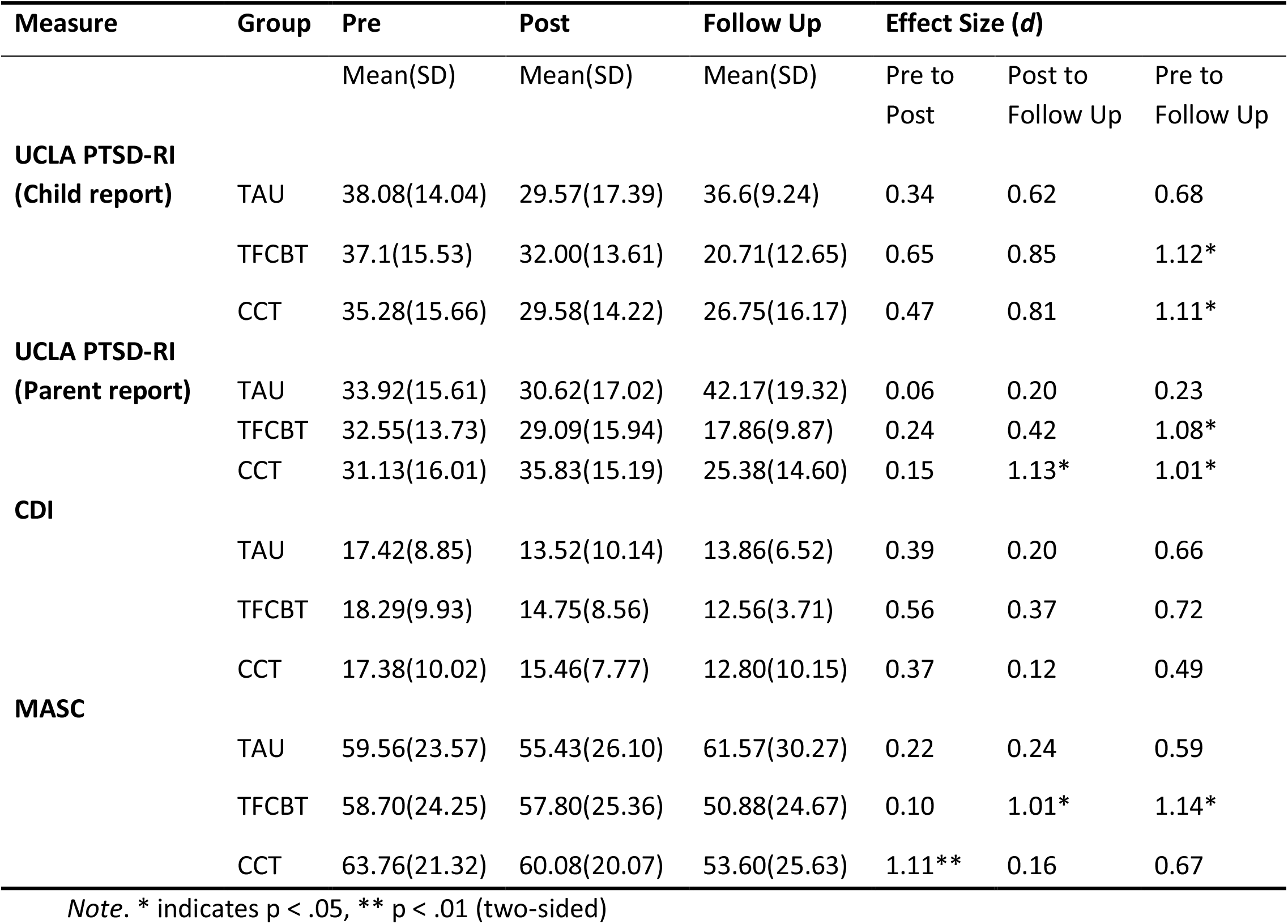
Means and Standard Deviations for Measures by Treatment Group

**Figure 1.**
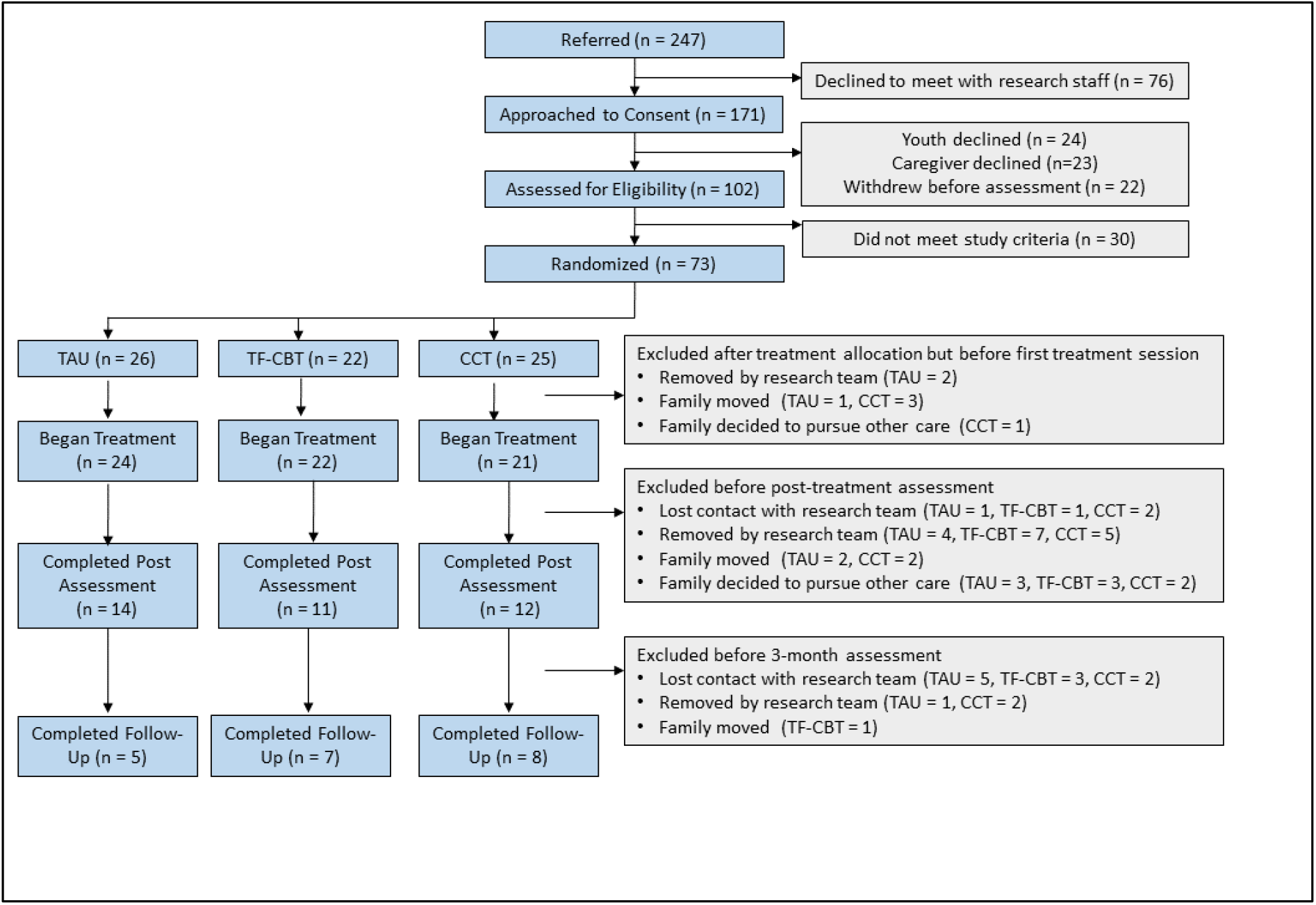
CONSORT diagram showing participant flow from referral through 3-month follow-up.

**Figure 2.**
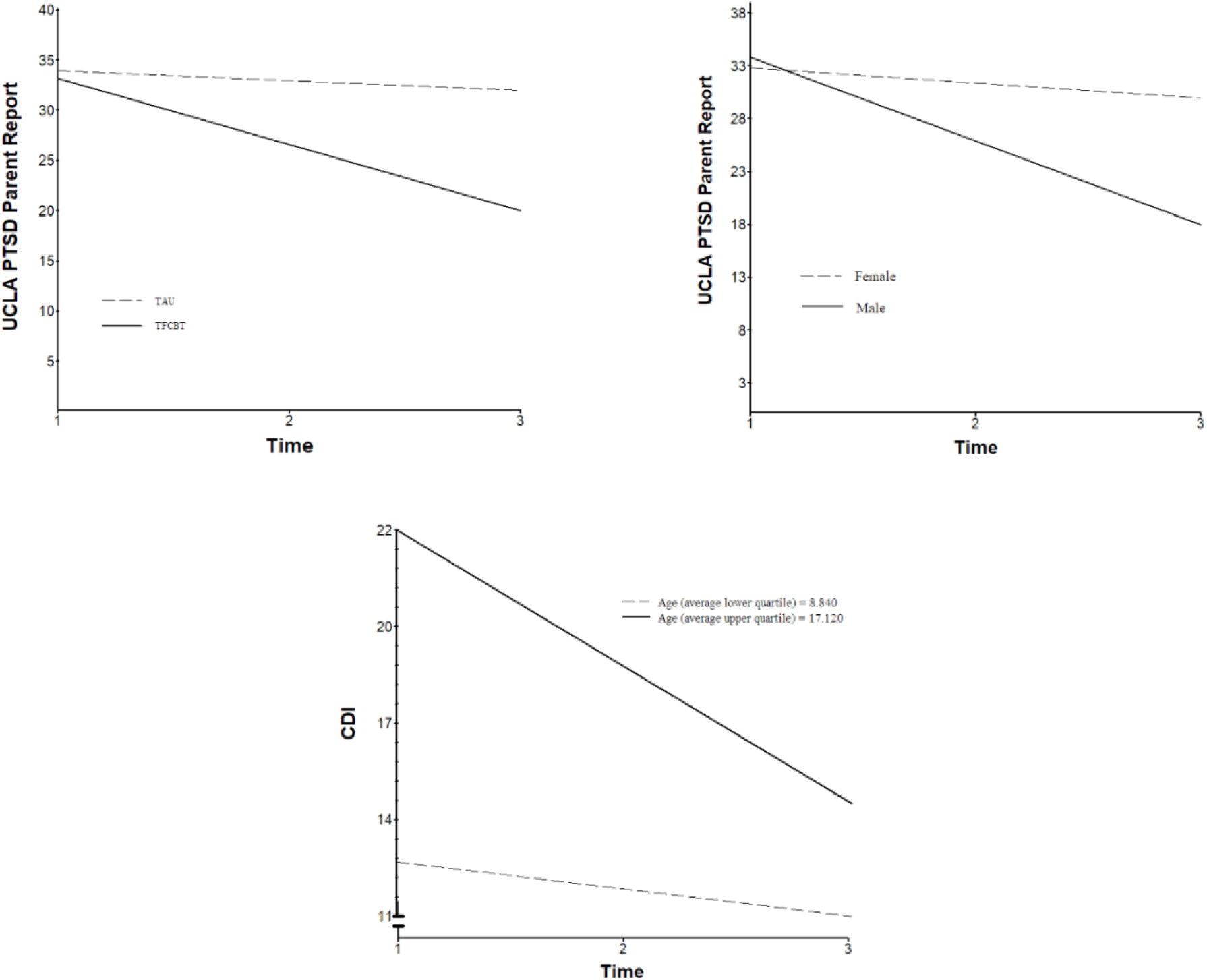
Significant and trending effects over time by treatment group, age, and gender

**Figure 3.**
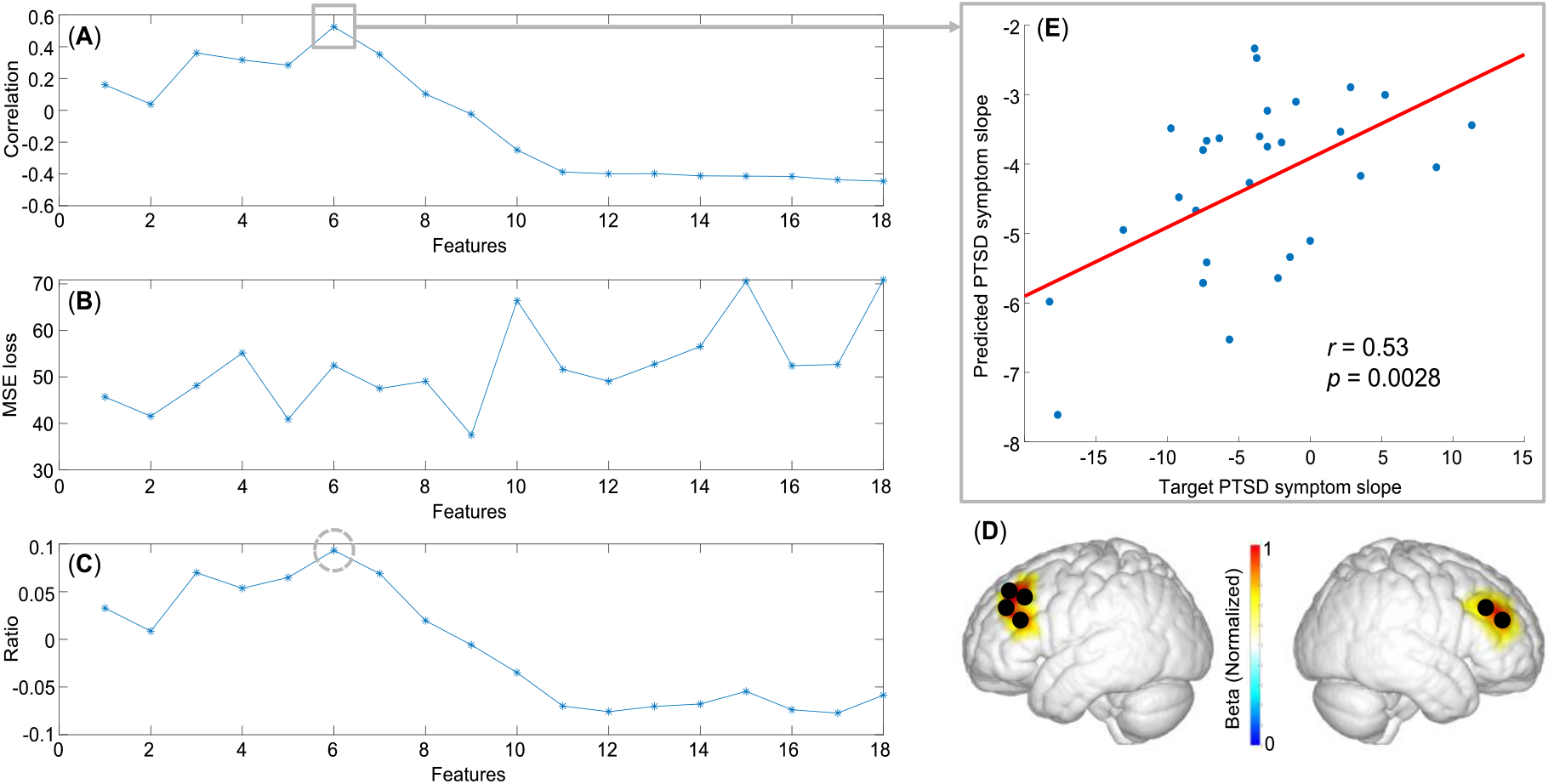
Training process of the predictive model across the different feature numbers are presented: (**A**) correlation between the predicted slope scores and the target slope score; (**B**) Mean Square Error (MSE) of the model; (**C**) ratio between correlation and MSE, with an optimal feature number of six (black dash circle). The six features (i.e., cortical responses from six fNIRS channels as depicted in **D** and the correlation with PTSD symptom improvement (*r* = 0.53, *p* < 0.0028) in **E**.

**Figure 4.**
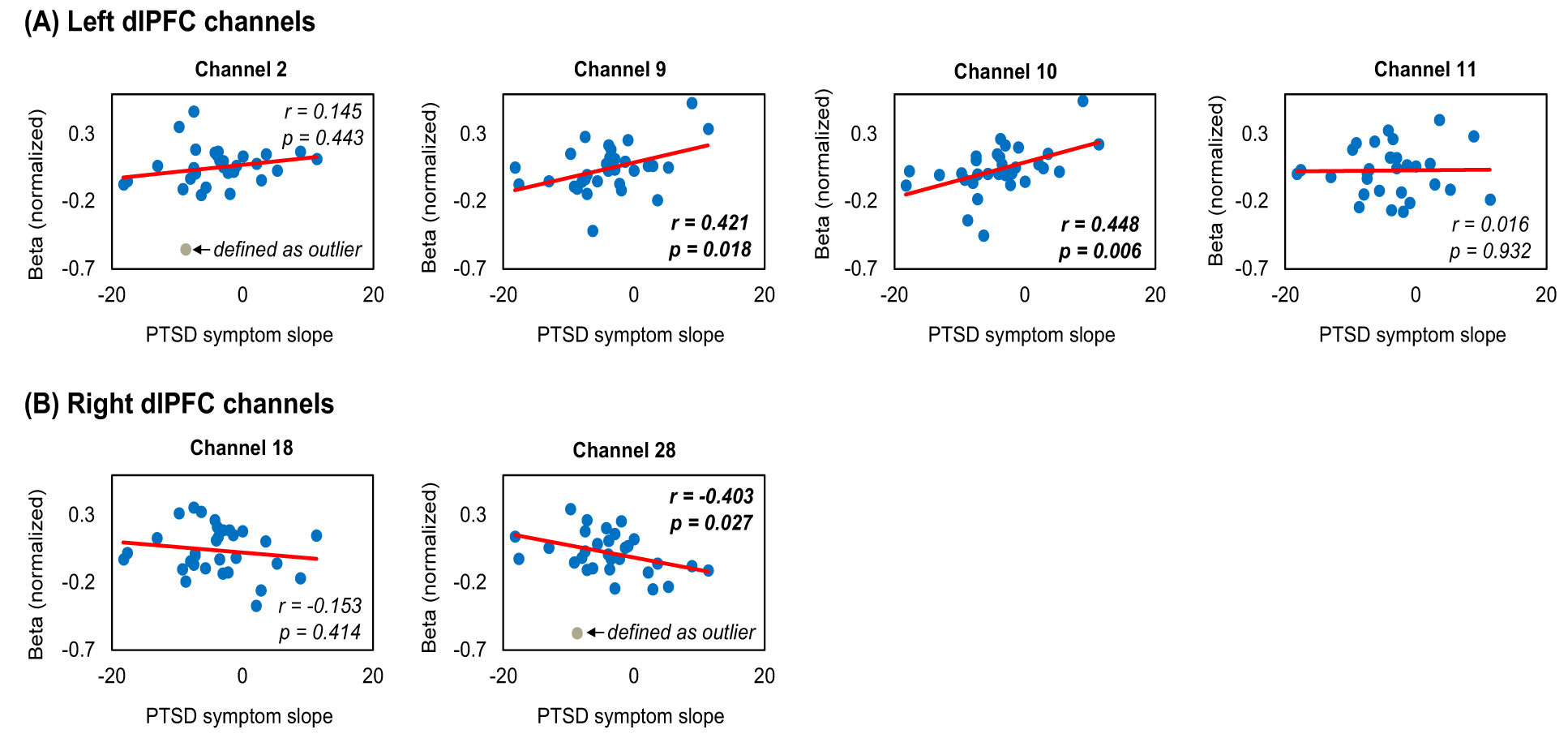
Associations between normalized beta values and PTSD symptom slope for each of the six fNIRS channels, including channels of the left dlPFC (**A**) and the right dlPFC (**B**).

## Notes

### Competing Interest Statement

The authors have declared no competing interest.

### Clinical Trial

NCT02926677

### Funding Statement

This work was supported by a gift from the Eucalyptus Foundation, San Francisco, USA.

### Author Declarations

This research was approved by the Stanford University Institutional Review Board and registered on clinicaltrials.gov (NCT02926677).

## References

Aebi, M., Mohler-Kuo, M., Barra, S., Schnyder, U., Maier, T., & Landolt, M.A. (2017). Posttraumatic stress and youth violence perpetration: A population-based cross-sectional study. European Psychiatry, 40, 88–95.

Alisic, E., Zalta, A.K., van Wesel, F., Larsen, S.E., Hafstad, G.S., Hassanpour, K., & Smid, G.E. (2014). Rates of post-traumatic stress disorder in trauma-exposed children and adolescents: meta-analysis. The British Journal of Psychiatry, 204(5), 335–340.

American Academy of Child and Adolescent Psychiatry (AACAP) (2010). Practice parameter for the assessment and treatment of children and adolescents with posttraumatic stress disorder. Journal of the American Academy of Child & Adolescent Psychiatry, 49, 414–30.

American Psychiatric Association. (2013). Diagnostic and statistical manual of mental disorders (5^th^ ed.). https://doi-org.ezproxy.frederick.edu/10.1176/appi.books.9780890425596

Balters, S., Li, R., Espil, F.M., Piccirilli, A., Liu, N., Gundran, A., Carrion, V.G., Weems, C.F., Cohen, J.A., and Reiss, A.L. (2021). “Functional near-infrared spectroscopy brain imaging predicts symptom severity in youth exposed to traumatic stress.” Journal of psychiatric research 144: 494–502.

Bryk, A. S., & Raudenbush, S. W. (1987). Application of hierarchical linear models to assessing change. Psychological bulletin, 101(1), 147.

Carrion VG, Kletter H, Weems CF, Berry RR, & Rettger JP (2013). Cue-centered treatment for youth exposed to interpersonal violence: a randomized controlled trial. Journal of Traumatic Stress, 26, 654–62.

Carrion, V.G., Garrett, A., Menon, V., Weems, C.F., Reiss, A.L. (2008). Posttraumatic stress symptoms and brain function during a response-inhibition task: An fMRI study in youth. Depression and Anxiety, 25(6), 514–526.

Carrion, V.G., Haas, B.W., Garrett, A., Song, S., & Reiss, A.L. (2010). Reduced hippocampal activity in youth with posttraumatic stress symptoms: An fMRI study in youth. Journal of Pediatric Psychology 35(5), 559–569.

Carrion, V. G., & Weems, C. F. (2017). Neuroscience of Pediatric PTSD. New York: Oxford University Press. ISBN: 9780190201968 https://global.oup.com/academic/product/neuroscience-of-pediatric-ptsd-9780190201968?cc=us&lang=en&

Carrion, V.G. (2016). Cue-centered therapy for youth experiencing posttraumatic symptoms: a structured multi-modal intervention, therapist guide. New York: Oxford University Press. Change. Psychological bulletin, 101(1), 147.

Chorpita, B. F., Daleiden, E. L., Park, A. L., Ward, A. M., Levy, M. C., Cromley, T., … & Krull, J. L. (2017). Child STEPs in California: A cluster randomized effectiveness trial comparing modular treatment with community implemented treatment for youth with anxiety, depression, conduct problems, or traumatic stress. Journal of consulting and clinical psychology, 85(1), 13.

Cisler, J. M., Sigel, B. A., Steele, J. S., Smitherman, S., Vanderzee, K., Pemberton, J., … & Kilts, C. D. (2016). Changes in functional connectivity of the amygdala during cognitive reappraisal predict symptom reduction during trauma-focused cognitive–behavioral therapy among adolescent girls with post-traumatic stress disorder. Psychological medicine, 46(14), 3013–3023.

Cohen, J.A., & Mannarino, A.P. (2008). Trauma-focused cognitive behavioural therapy for children and parents. Child Adolescent Mental Health, 13, 158–62.

Cohen, J.A., Mannarino, A.P., Kliethermes, M., & Murray, L.A. (2012). Trauma-focused CBT for youth with complex trauma. Child Abuse & Neglect, 36(6), 528–41.

Cohen, JA, Mannarino, A. & Deblinger, E (2017) Treating trauma and traumatic grief in children & adolescents, 2^nd^ edition. New York: Guilford Press.

Copeland-Linder, N. (2008). Posttraumatic stress disorder. Pediatr Rev. 2008;29:103–104.

Copeland, W.E., Keeler, G., Angold, A., & Costello, E.J. (2007). Traumatic events and posttraumatic stress in childhood. Archives of General Psychiatry, 64(5), 577–584.

Cui, X., Bray, S. and Reiss, A.L. (2010). Functional near infrared spectroscopy (NIRS) signal improvement based on negative correlation between oxygenated and deoxygenated hemoglobin dynamics. Neuroimage, 49(4), pp.3039–3046.

De Haan, A. M., Boon, A. E., De Jong, J. T., Hoeve, M., & Vermeiren, R. R. (2013). A meta-analytic review on treatment dropout in child and adolescent outpatient mental health care. Clinical psychology review, 33(5), 698–711.

Etkin, A., Egner, T. & Kalisch, R. (2011). Emotional processing in anterior cingulate and medial prefrontal cortex. Trends Cogn. Sci. 15, 85–93.

Ferrari, M. and Quaresima, V., (2012). A brief review on the history of human functional near-infrared spectroscopy (fNIRS) development and fields of application. Neuroimage, 63(2), pp.921–935.

Ganella, D.E., Barendse, M.E., Kim, J.H., Whittle, S. (2017). Prefrontal-amygdala connectivity and state anxiety during fear extinction recall in adolescents. Front. Hum. Neurosci. 11, 587.

Garrett, A., Cohen, J. A., Zack, S., Carrion, V., Jo, B., Blader, J., … & Agras, W. S. (2019). Longitudinal changes in brain function associated with symptom improvement in youth with PTSD. Journal of psychiatric research, 114, 161–169.

Garrett, A.S., Carrion, V.G., Kletter, H., Karchemskiy, A., Weems, C.F., & Reiss, A. (2012). Brain activation to facial expressions in youth with PTSD symptoms. Depression and Anxiety, 29(5), 449–459.

Gee, D.G. (2016). Sensitive periods of emotion regulation: influences of parental care on frontoamygdala circuitry and plasticity.

Giaconia, R.M., Reinherz, H.Z., Silverman, A.B., Pakiz, B., Frost, A.K., & Cohen, E. (1995). Traumas and posttraumatic stress disorder in a community population of older adolescents. Journal of the American Academy of Child & Adolescent Psychiatry, 34(10), 1369–80.

Herringa, R. J. (2017). Trauma, PTSD, and the developing brain. Current psychiatry reports, 19(10), 1–9.

Keding, T. J., & Herringa, R. J. (2016). Paradoxical prefrontal–amygdala recruitment to angry and happy expressions in pediatric posttraumatic stress disorder. Neuropsychopharmacology, 41(12), 2903–2912.

Koch, S. B. et al. (2016). Aberrant resting-state brain activity in posttraumatic stress disorder: A meta-analysis and systematic review. Depress. Anxiety 33, 592–605. 64.

Kondo, H., Osaka, N., & Osaka, M. (2004). Cooperation of the anterior cingulate cortex and dorsolateral prefrontal cortex for attention shifting. Neuroimage, 23(2), 670–679.

Kovacs, M. (2003). Children’s depression inventory (CDI). Toronto: Multi-Health System.

Lanius, R.A., Williamson, P.C., Bluhm, R.L., Densmore, M., Boksman, K., Neufeld, R.W., Gati, J.S., Menon, R.S., (2005). Functional connectivity of dissociative responses in posttraumatic stress disorder: a functional magnetic resonance imaging investigation. Biol. Psychiatr. 57 (8), 873–884.

Lanius, R.A., Williamson, P.C., Boksman, K., Densmore, M., Gupta, M., Neufeld, R.W., Gati, J.S., Menon, R.S., 2002. Brain activation during script-driven imagery induced dissociative responses in PTSD: a functional magnetic resonance imaging investigation. Biol. Psychiatr. 52 (4), 305–311.

Lenz, A. S., & Hollenbaugh, K. M. (2015). Meta-analysis of trauma-focused cognitive behavioral therapy for treating PTSD and co-occurring depression among children and adolescents. Counseling Outcome Research and Evaluation, 6(1), 18–32.

Lorch, R. F., & Myers, J. L. (1990). Regression analyses of repeated measures data in cognitive research. Journal of Experimental Psychology: Learning, Memory, and Cognition, 16(1), 149–157. doi: 10.1037/0278-7393.16.1.149

Macdonald, A., Danielson, C.K., Resnick, H.S., Saunders, B.E., & Kilpatrick, D.G. (2010). PTSD and comorbid disorders in a representative sample of adolescents: The risk associated with multiple exposures to potentially traumatic events. Child Abuse & Neglect, 34(10), 773–783.

Malarbi, S., Abu-Rayya, H.M., Muscara, F., & Stargatt, R. (2017). Neuropsychological functioning of childhood trauma and post-traumatic stress disorder: A meta-analysis. Neuroscience & Biobehavioral Reviews, 72, 68–86.

March, J. S., Parker, J. D. A., Sullivan, K., Stallings, P., & Conners, C. (1997). The Multidimensional Anxiety Scale for Children (MASC): Factor structure, reliability, and validity. Journal of the American Academy of Child & Adolescent Psychiatry, 36, 554–565

Matlow, R. (2019). General principles of psychotherapy. In: Carrion V, editor. Assessing and treating youth exposed to traumatic stress. Washington DC: American Psychiatric Association Publishing.

McCrimmon, A. W., & Smith, A. D. (2013). Review of the Wechsler abbreviated scale of intelligence, (WASI-II).

Miller, L. N., Simmons, J. G., Whittle, S., Forbes, D., & Felmingham, K. (2021). The impact of posttraumatic stress disorder on event-related potentials in affective and non-affective paradigms: A systematic review with meta-analysis. Neuroscience & Biobehavioral Reviews, 122, 120–142.

Molavi, B. and Dumont, G.A., (2012). Wavelet-based motion artifact removal for functional near-infrared spectroscopy. Physiological measurement, 33(2), p.259.

Pfister, R., Schwarz, K., Carson, R., & Jancyzk, M. (2013). Easy methods for extracting individual regression slopes: Comparing SPSS, R, and Excel. Tutorials in Quantitative Methods for Psychology, 9(2), 72–78.

Plichta, M.M., Herrmann, M.J., Baehne, C.G., Ehlis, A.C., Richter, M.M., Pauli, P. and Fallgatter, A.J. (2006). Event-related functional near-infrared spectroscopy (fNIRS): are the measurements reliable?. Neuroimage, 31(1), pp.116–124.

Raudenbush, S. W. (2004). HLM 6: Hierarchical linear and nonlinear modeling. Scientific Software International.

Russell, J. D., Neill, E. L., Carrión, V. G., & Weems, C. F. (2017). The network structure of posttraumatic stress symptoms in children and adolescents exposed to disasters. Journal of the American Academy of Child & Adolescent Psychiatry, 56(8), 669–677.

Sanchez-Lopez, A., Vanderhasselt, M. A., Allaert, J., Baeken, C., & De Raedt, R. (2018). Neurocognitive mechanisms behind emotional attention: Inverse effects of anodal tDCS over the left and right DLPFC on gaze disengagement from emotional faces. Cognitive, Affective, & Behavioral Neuroscience, 18(3), 485–494.

Sanchez, A., Vanderhasselt, M. A., Baeken, C., & De Raedt, R. (2016). Effects of tDCS over the right DLPFC on attentional disengagement from positive and negative faces: an eye-tracking study. Cognitive, Affective, & Behavioral Neuroscience, 16(6), 1027–1038.

Santosa, H., Zhai, X., Fishburn, F. and Huppert, T. (2018). The NIRS brain AnalyzIR toolbox. Algorithms, 11(5), p.73.

Schwartz, B., Kaminer, D., Hardy, A., Nöthling, J., & Seedat, S. (2021). Gender differences in the violence exposure types that predict PTSD and depression in adolescents. Journal of interpersonal violence, 36(17-18), 8358–8381.

Shonkoff, JP, Garner, AS, Siegel, BS, Dobbins, MI, Earls, MF, McGuinn, L, … & Committee on Early Childhood, Adoption, and Dependent Care. (2012). The lifelong effects of early childhood adversity and toxic stress. Pediatrics, 129(1), e232–e246.

Sitarenios, G., & Stein, S. (2004). Use of the Children’s Depression Inventory. Software International.

Sripada, R. K. et al. (2012). Altered resting-state amygdala functional connectivity in men with posttraumatic stress disorder. J. Psychiatry Neurosci. JPN 37, 241.

Wechsler, D. (2011). Wechsler Abbreviated Scale of Intelligence—Second Edition (WASI-II). San Antonio, TX: NCS Pearson.

Weems, C.F., & Neill, E.L. (2020). Empirically supported treatment options for children and adolescents with posttraumatic stress disorder: Integrating network models and treatment components. Current Treatment Options in Psychiatry (7), 103–119.

Weems, C.F., Russell, J.D., Herringa, R.J., Carrion, V.G. (2021). Translating the neuroscience of adverse power analysis program for the social, behavioral, and biomedical sciences. American Psychologist, 76(2):188.

Weems, C. F., Russell, J. D., Neill, E. L., & McCurdy, B. H. (2019). Annual research review: Pediatric posttraumatic stress disorder from a neurodevelopmental network perspective. Journal of Child Psychology and Psychiatry, 60(4), 395–408.

Wei, C., Hoff, A., Villabø, M. A., Peterman, J., Kendall, P. C., Piacentini, J., … & March, J. (2014). Assessing anxiety in youth with the multidimensional anxiety scale for children. Journal of Clinical Child & Adolescent Psychology, 43(4), 566–578.

Weissman, M. M., Verdeli, H., Gameroff, M. J., Bledsoe, S. E., Betts, K., Mufson, L., … Wickramaratne, P. (2006). National survey of psychotherapy training in psychiatry, psychology, and social work. Archives of General Psychiatry, 63(8), 925–934.

Weisz, J. R., Kuppens, S., Eckshtain, D., Ugueto, A. M., Hawley, K. M., & Jensen-Doss, A. (2013). Performance of evidence-based youth psychotherapies compared with usual clinical care: A multilevel meta-analysis. JAMA psychiatry, 70(7), 750–761.

Weisz, J. R., Bearman, S. K., Ugueto, A. M., Herren, J. A., Evans, S. C., Cheron, D. M., … & Jensen-Doss, A. (2020). Testing robustness of child STEPs effects with children and adolescents: A randomized controlled effectiveness trial. Journal of Clinical Child & Adolescent Psychology, 49(6), 883–896.

Wyatt, J.S., Delpy, D.T., Cope, M., Wray, S. and Reynolds, E.O.R., (1986). Quantification of cerebral oxygenation and haemodynamics in sick newborn infants by near infrared spectrophotometry. The Lancet, 328(8515), pp.1063–1066.

Yoon, S., Steigerwald, S., Holmes, M.R., & Perzynski, A.T. (2016). Children’s Exposure to Violence: The Underlying Effect of Posttraumatic Stress Symptoms on Behavior Problems: Child Exposure to Violence. Journal of Traumatic Stress, 29(1), 72–79.

